# Diagnostic value of self-report compared with Beck Depression Inventory (BDI-II) in screening of depression

**DOI:** 10.1101/2020.04.29.20085852

**Authors:** Mehran zarghami, Fatemeh Taghizadeh, Mahmood mousazadeh

**Affiliations:** Department of Psychiatry, School of Medicine, Psychiatry and Behavioral Sciences Research Center, Addiction Institute, Mazandaran University of Medical Sciences, Sari, Iran; Department of Epidemiology, Faculty of Medicine, Mazandaran University of Medical Sciences, Sari, Iran

**Keywords:** Depression, Beck depression inventory, Self-reporting

## Abstract

**Background:** Depression is a common cause of mortality and morbidity worldwide. To detect depression, we compared Beck Depression Inventory scoring as a valid tool with participants self-reporting depression.

**Methodology:** This cross-sectional study aimed to explore the diagnostic values of self-reporting in patients’ with depression comparing to Beck Depression Inventory scoring in Mazandaran Persian cohort study, with a total of 1300 samples. The sample size was determined to include 155 participants through the census method. In order to increase the test power, 310 healthy participants were included in the study through random selection. In order to evaluate the diagnostic value of self-reporting, BDI-II was completed by blind interviewing to the case group as well as to another group who reported that they were not depressed, as control.

**Results:** Sensitivity, specificity, accuracy, false positive, false negative, positive and negative predictive values of self-reporting was calculated 58.4%, 79.1%,73.4%, 20.8%, 41.6%, 51.8%, and 83.2% for the total population respectively, as well as, sensitivity, specificity, accuracy, positive and negative predictive values of self-report in males were 83.3%, 77.2%, 77.1%, 43.8% and 95.6% and 53.7%, 78.1%, 71.2%, 49.2%, and 81.1% for females, respectively.

**Conclusion:** The positive predictive value and sensitivity of self-reporting are insufficient in total population and females, and therefore self-reporting cannot detect depressed patients, but regarding to its average positive predictive value, perhaps, it can be used to identify non-depressant individuals.

## Introduction

The World Health Organization (WHO) has identified depression as the fourth reason of disability in the world, accounting for the greater portion of non-lethal diseases, and predicts it to be the second cause of death by 2020[1-3]. In a review study, the prevalence of lifetime depression varied from 1.5 percent in Taiwan to 19 percent in Lebanon. The average in western Germany was 9.2 percent, and in Edmonton in Canada, it was reported at 9.6 percent [1]. An international research by the WHO, reported the prevalence of major depression in the general population to be from 1 percent in the Czech Republic to 16.9 percent in the United States, with an average of 8.3 percent in Canada, and up to 9 percent in Chile [1]. The average global prevalence of depression is reported to be about 15 percent [4]. The prevalence of depression in the Iranian adult population is assessed at 21 percent [4]. Regarding the high importance of this disorder, screening of this serious condition and timely management would be an important subject. There are several assessments for diagnosis of depression, namely Hamilton Depression Rating Scale (HAM-D), Zung Self-Rating Depression Scale [5], Montgomery-Asberg Depression Rating Scale, HADS [6], Geriatric Depression Scale, and the General Health Questionnaire (GHQ). They have few items for depression, except the HAM-D [4], these depression assessment tools were developed as a measure of treatment outcome rather than a diagnostic or screening depression [7]. However, the Beck Depression Inventory (BDI) assesses both the psychosomatic and the physical symptoms, and its effectiveness has been discussed in many studies[7,4]. This tool has been used in more than 7,000 researches so far. The theoretical assumption of the BDI relied upon the negative believes that distorted cognition is the core of depression characteristic [8]. This inventory is a valuable instrument, with high reliability to discriminate depressed and non-depressed participants, and its content, structural and concurrent validity has been approved[8]. This tool has been revised two times and the latest version (BDI-II) was published in 1996 [9]. The available psychometric evidence showed that the BDI-II could be noticed as a valid cost-effective inventory for measuring the depression severity, with wide applicability for research and clinical practice [8].

BDI-II is the approved screening tool for assessment of depression in the Persian cohort in Mazandaran, Iran. As in some studies, it has been indicated that the prevalence of depression measured through diagnostic scales by patients has been higher than the diagnostic interview and self-report results[8], the researchers decided to compare the diagnostic value of the depression with BDI-II in Mazandaran’s Persian cohort study with self-reported depression. The authors predicted that self-reporting would provide important diagnostic information when applied to patients with depression.

## Methodology

In this cross-sectional study, we used a subset of data collected in Tabari cohort (Mazandaran’s Persian cohort study), which is part of the national cohort, entitled as Prospective Epidemiological Research in Iran (Persian)[10, 11]. For conducting this study, 1300 participants, aged 35–70 years living in urban areas of Sari, Mazandaran, Iran, were enrolled. As part of data collection in Tabari cohort, a standardized questionnaire consisting of general information, socioeconomic status, occupational history, type of fuel used, characteristics of the habitat, life style, history of fertility, history of chronic diseases, drug use, familial history of diseases, oral health, physical examination, physical disabilities, sleep status, physical activity and smoking and drinking were completed. All the participants were asked a question" Are you depressed? “. Among all the participants, 155 cases had a history of depression, which were selected as the case group. Among the remaining participants who did not report depression, 310 individuals were selected as control group randomly and matched in age and sex.

In order to evaluate the diagnostic value of self-reporting, BDI-II was completed to the case group as well as to another group who reported that they were not depressed.

Trained interviewers who were blind to the interviewees, dispatched to the households based on their Household Registry Number addresses to fill out the demographic questionnaire and the BDI-II. For illiterate participants, the questions were read and they answered without any elaborations or comments.

This cross-sectional study aimed to investigate the diagnostic values of self-reporting in patients with depression compared to BDI-II in Mazandaran’s Persian cohort study with a total of 1300 samples. The sample size was determined to include 116 participants based on the results of Kim et al. study [3], where the correct classification is reported to be 82 percent, with a confidence level of 95% and an accuracy of 0.07. With the effect size equal to 1.3 times, the sample size was estimated 155 participants that allocated through the census method. In order to increase the test power by 2 times, 310 healthy participants (by self-reporting) were entered the study through random selection (based on the available list) and the following formula:

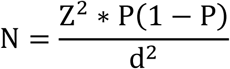

## Statistical analysis

Data was entered into SPSS (version 22) software for statistical analysis. After filtering, the distribution of characteristics of the studied population was presented through descriptive tests such as frequency, mean, and standard deviation. Also, sensitivity, specificity, positive and negative predictive value and accuracy of self-report method were determined.

## Questionnaires

### 1. Demographic information Questionnaire

This questionnaire included demographic information such as age, sex, and history of depression.

### 2. The Beck Depression Inventory (BDI-II)

The BDI-II is a multiple-choice self-report inventory, consisting of 21 questions, first developed by Aaron Beck in 1961 [12]. The 21 items are based on symptoms follow as:

1. Sadness 2. Pessimism 3. Sense of failure 4. Lack of satisfaction 5. Guilt 6. Feelings of being punished 7. Self-hate, 8. Self-accusations, 9. Self-harm 10. Crying spells (crying periods), 11. Early suffering (excitability), 12. Social isolation, 13. Undecidedness, 14. Self-thought (change in body image), 15. Weakness and slowness (slowness in doing a task, slowness at work), 16. Sleep disturbance (insomnia), 17. Fatigue 18, decreased appetite (loss of appetite), 19. Weight loss, 20. Somatic preoccupation, 21. Loss of libido (12, 13).

In this inventory, 4 to 6 questions are asked concerning each of the mentioned items based on one of the symptoms of the illness, ranging from the mildest to the most severe aspect of the mentioned attribute [13].The quantitative values of each item from 0 to 3 are determined as mild to severe disorder. Several forms of this questionnaire have been prepared. Here the regular form includes 21 items [13].This questionnaire is a self-assessment instrument and takes 5 to 10 minutes to complete.

### Scoring

The total score ranges from 0 to 63. These marks are interpreted in the diagnosis of depression as follows: normal (no clinical disease (1–13), mild depression (14 to 19), moderate depression (20 to 28), and severe depression (29 to 63) [13].

It should be noted that, even though this inventory was designed for use in clinical populations; besides, it could also be used in normal populations [13].

### Reliability and validity

Beck, Stier, and Garbin obtained the internal consistency coefficients at 0.73 to 0.92, with an average of 0.86 [14]. The content of the BDI materials included six of the nine categories of DSM-III for diagnosis of depression [2].The correlation of this test with the Hamilton scale for depression (0.73), Zong’s depression scale (0.76), and MMPI depression scale (0.76) were obtained [14]. The correlation coefficient was obtained as 0.54 through the MMPI Depression Scale [15]. However, factor analysis showed a robust dimension of general depression composed by two constructs: cognitive-affective and somatic-vegetative [8].These data support the reliability and concurrent validity of the BDI-II-Persian as a measure of depressive symptoms in nonclinical samples [16].

### Cut-off of BDI-II

The cut-off score for screening of depression varied according to the type of sample. In a study in Iran, the best BDI-II cut-off was 14, with sensitivity of 62% (95% CI (43%, 81%)), specificity of 81% (95% CI (72%, 90%)), PPV of 53%, NPV of 85% [16]. The internal consistency was described as around 0.9 and the test-retest reliability ranging from 0.73 to 0.96[8].Accordingly, in this study, a score of 14 was considered as the cut-off point for screening of depression.

## Results

### 1. Biographic characteristics of population

141 (32%) of the participates were cases, and 310 (68%) of them were controls completed the study (Table 1). Of all the participants, 69 (15%) were male and 382 (85%) female, 437(96%) married, 10 (2%) widowed, 3 single, and 1 divorced. With regard to age, 136 (30%) of the participants were 37–46 years old, 178 (39%) 47–56 years old, 117 (25%) 57–66 years old, and 20 (4%) 66–72 years old.

**Table 1.** Frequency of population characteristics (sex, age, being under depression treatment, existence of depression in first grade relatives) in the case and control groups based on self-report

| **Group** | **Case**<br>**F (%)** | **Control**<br>**F (%)** | **Total**<br>**F (%)** |
| --- | --- | --- | --- |
| **Male** | 23(16) | 46(15) | 69(15) |
| **Female** | 118(84) | 264(85) | 382(85) |
| **Total** | 141(100) | 310(100) | 451(100) |
| **age group** | **Case**<br>**F (%)** | **Control**<br>**F (%)** | **Total**<br>**F (%)** |
| **35-50** | 57(41) | 125(41) | 182(40) |
| **51-72** | 84(59) | 185(59) | 269(59) |
| **Total** | 141(100) | 310(100) | 451(100) |

### 2. Sensitivity, specificity, positive and negative predictive values

With the cut-off 14 of BDI-II, sensitivity, specificity, accuracy, false positive, false negative, positive and negative predictive values of self-reporting were calculated 58.4%, 79.1%,73.4%, 20.8%, 41.6%, 51.8%, and 83.2% for the total population respectively.

Sensitivity, specificity, accuracy, positive and negative predictive values of self-report in males were 83.3%, 77.2%, 77.1%, 43.8% and 95.6% and 53.7%, 78.1%, 71.2%, 49.2%, and 81.1% for females respectively.

In addition, sensitivity, specificity, accuracy, positive and negative predictive values of self-report was 79.2%, 75.9%, 76.4%, 33.3%, and 96% for the 35–50 age group, and 51%, 79.8%, 69.5%, 58.3%, and 74.6% for the 51–72 age group respectively.

In addition, Table 1 shows the frequency of population characteristics in the case and control groups based on self-report; moreover, Table 2 shows the frequency of depression according to BDI-II (sex, age, depression, depression in family) in the case and control groups according to BDI-II, respectively. Table 3 presents the frequency of depression in the case and control groups based on BDI-II

**Table 2.** Distribution of sex, age, family history of depression, being depressed according to BDI-II, and being under treatment of depression

| **Gender** | **Without**<br>**depression**<br>**F (%)** | **Depressed**<br>**F (%)** | **Total**<br>**F (%)** |
| --- | --- | --- | --- |
| **Male** | 57(18) | 12(10) | 69(15) |
| **Female** | 274(82) | 108(90) | 382(84) |
| **Total**<br>**F (%)** | 331(100) | 120(100) | 451(100) |
| **age group** | **Without**<br>**depression**<br>**F (%)** | **Depressed**<br>**F (%)** | **Total**<br>**F (%)** |
| **35-50** | 158(47) | 24(20) | 182(40) |
| **51-72** | 173(52) | 96(80) | 269(59) |
| **Total**<br>**F (%)** | 331(100) | 120(100) | 451(100) |
| **being under**<br>**depression**<br>**treatment** | **Without**<br>**depression**<br>**F (%)** | **Depressed**<br>**F (%)** | **Total**<br>**F (%)** |
| **No** | 273(82) | 57(47) | 330(73) |
| **Yes** | 58(17) | 63(52) | 121(26) |
| **Total**<br>**F (%)** | 331(100) | 120(100) | 451(100) |
| **Family history of**<br>**depression** | **Without**<br>**depression**<br>**F (%)** | **Depressed**<br>**F (%)** | **Total**<br>**F (%)** |
| **No** | 298(90) | 107(89) | 405(89) |
| **Yes** | 33(10) | 13(10) | 46(10) |
| **Total**<br>**F (%)** | 330(100) | 120(100) | 451(100) |

**Table 3.** Frequency of depression in the case and control groups based on BDI-II

| **Group** | **Without**<br>**depression**<br>**F (%)** | **Mild**<br>**F (%)** | **Moderate**<br>**F (%)** | **Severe**<br>**F (%)** | **Total**<br>**F (%)** |
| --- | --- | --- | --- | --- | --- |
| Case | 73(52) | 34(24) | 28(20) | 6(4) | **141(100)** |
| Control | 258(84) | 27(8) | 22(7) | 3(1) | **310(100)** |
| Total | 331(74) | 61(13) | 50(11) | 9(1) | **451(100)** |

## Discussion

To our knowledge, this is the first study to compare the prevalence of depression with self-reporting and BDI-II as well as the first study to evaluate depression screening in a general population with the patients’ self-report. In this study, the prevalence of depression was assessed blindly (being case or control) using BDI-II in two groups. According to the results, the sensitivity and specificity of self-reporting was found to be low, with many of the cases being found not depressed via BDI-II (Table 3). It was concluded that self-reporting was not suitable for screening for depression in this population, and thus, there is a need to use a scale such as BDI-II as the gold standard for depression screening.

Individual clinical interview is the "gold standard" for diagnosis of depression[17]. However, this approach may be problematic for screening of depression in large populations. In the Persian cohort study, the participants were asked only one question in this case, namely ‘Are you depressed based on physician’s opinion?’ This study aimed to evaluate the diagnostic value of self-reporting compared with one of the most popular scales for depression screening.

The BDI is one of the most well-known tools for screening of depression in general population and psychiatric patients [17, 18]. One of the problems of the BDI is that it did not completely include all of the symptoms in the DSM in depression criteria [19]. This revised instrument does not rely on any certain theory of depression [17]. The **BDI**-II has a good reliability and validity (21). The correlation between BDI-II and BDI-I has been described strong [20].

The correlation between BDI-II and BDI-I has been reported high[8] With respect to the Multiscale Depression Inventory as a ‘gold standard’, the curve of receiver operational characteristic showed **BDI**-II to be an adequate diagnostic measure and that the optimal total cutoff score was 18.5 [20] With this cut-off score, 25% of multiple sclerosis patients were positively identified as having clinically relevant depression. The result of this study showed that the **BDI**-II is a valid, reliable, and simple tool for depression detecting and grading [20]. In a systematic review study on psychometric BDI-II characteristics, 118 study were assigned into three groups: non-clinical, medical participants, psychiatric or institutionalized participants. The internal consistency was obtained 0.9 and the test-retest reliability was revealed 0.73 to 0.96. The cut-off score for depression screening varied according to the variety of participants. Factor analysis presented a strong dimension of general depression composition with 2 constructs: somatic-vegetative and cognitive-affective[8].The **BDI**-II is a valid psychometric instrument, showing high reliability, capacity to discriminate between depressed and non-depressed subjects, and improved concurrent, content, and structural validity. Based on the available psychometric evidence, the **BDI**-II can be viewed as a cost-effective instrument for measuring the severity of **depression**, with broad applicability for research and clinical practice worldwide [8]. This questionnaire was used in our study for depression screening in the general population.

Concerning the self-report in the research, in a study, self-reported alcohol use was compared to biomarker tests via the Audit and 90-day recall for 193 women from prenatal clinics. The Audit was positive in 67.9% of the participants, and 65.3% of them directly reported drinking. Individual biomarkers revealed less drinking than self-reporting, but 64.8% had drinking-positive values on biomarkers, which were not different significantly from self-report. The biomarkers showed that 3.1% - 6.8% of participants lied about their drinking. The combined biomarker sensitivity was 95% - 80% and the specificity was 49% −76% for drinking in the 7 to 90 days ago. The best yield combined biomarker results was 89.6% with accuracy of 78.8% when evaluating 90 day drinking [21]. In confirmation of the conclusion of this study, many of the patients may have given contradictory answers or lied in self-report regarding their depression, or otherwise gave vague or ambiguous answers.

Another study evaluated the interactions among three selected FKBP5 single-nucleotide polymorphisms and objectively recorded ELS and self-reported early life stress (ELS) related to depression symptoms in midlife. The participants completed the Beck Depression Inventory at ages of 61.5 years (time 1) and 63.4 years (time 2); 165 and 181 participants were separated from their parents in childhood as a result of evacuations during World War II as indicated by self-reports and the Finnish National Archives registry, respectively. The relationship between objectively recorded ELS and self-reporting, and the average BDI score (mean of time 1 and time 2) or mild to severe **BDI** scores, or both, were moderated by the FKBP5 variants. FKBP5 variations combined with and objectively recorded ELS and self-reporting could predict more noticeable depression symptoms in midlife[22].

Moreover, in South Africa, 5059 participants aged above 40 years were entered in a study from 2014 to 2015. HIV biomarker testing, self-reporting HIV status and dried bloodspots were found during interviews at home. Regarding the biomarker results, 50.9% of participants reported knowing their HIV status and reported that accurately. PPV of self-reporting was 94.1%, NPV was 87.2 %, specificity of 99 % and sensitivity of 51.2 %. The patients on ART were more likely to reporting their HIV positive status, and the patients that reporting false-negatives were more likely to have older HIV tests. False-negative reports were mostly explained by lack of the testing, suggesting to be retreating HIV stigma in this setting [23]. It seems that drinking alcohol and HIV infection may be reflected as a stigma which can predict high rate of negative self-report. Concerning the results of this study, the stigma of having psychiatric disorders, such as depression, is a barrier to self-reporting of these problems, and a valid and reliable instrument is required to be arranged and conducted for detecting depression. In addition, sensitivity in our study was low by self-reporting compared to BDI-II as a gold standard.

## Conclusion

The positive predictive value and sensitivity of self-reporting are low, and therefore self-reporting cannot help in detecting depressed patients; however, concerning its average positive predictive value, perhaps, it can be used to identify non-depressant people.

## Data Availability

data is available when request

## Ethical approval

This study was approved by the Mazandaran University of Medical sciences’ ethics committee (IR.MAZUMS.REC.95.2V*7).

## References

[1] Kessler RC, Bromet EJ. The epidemiology of depression across cultures. Annual review of public health 2013;34:119.

[2] Beck AT, Ward C, Mendelson M. Beck depression inventory (BDI). Arch Gen Psychiatry 1961;4:561–71.

[3] Kim MH, Mazenga AC, Devandra A, Ahmed S, Kazembe PN, Yu X, et al. Prevalence of depression and validation of the Beck Depression Inventory-II and the Children’s Depression Inventory-Short amongst HIV-positive adolescents in Malawi. Journal of the International AIDS Society 2014;17.

[4] Noorbala A, Yazdi SB, Yasamy M, Mohammad K. Mental health survey of the adult population in Iran. The British Journal of Psychiatry 2004;184:70–3.

[5] Taylor R, Lovibond PF, Nicholas MK, Cayley C, Wilson PH. The utility of somatic items in the assessment of depression in patients with chronic pain: a comparison of the Zung Self-Rating Depression Scale and the Depression Anxiety Stress Scales in chronic pain and clinical and community samples. The Clinical journal of pain 2005;21:91–100.

[6] Rahimian Boogar I TS, Nikaeen N. The Comparison of the Resiliency and Psychological Risk Factors between the Youth of Smokers and Non-smokers JRUMS 2015;13:655–68.

[7] Pop-Jordanova N. BDI in the Assessment of Depression in Different Medical Conditions. Prilozi (Makedonska akademija na naukite i umetnostite Oddelenie za medicinski nauki) 2017;38:103–11.

[8] Wang YP, Gorenstein C. Psychometric properties of the Beck Depression Inventory-II: a comprehensive review. Revista brasileira de psiquiatria (Sao Paulo, Brazil: 1999) 2013;35:416–31.

[9] Beck AT, Steer RA, Ball R, Ranieri W. Comparison of Beck Depression Inventories -IA and -II in psychiatric outpatients. Journal of personality assessment 1996;67:588–97.

[10] Poustchi H, Eghtesad S, Kamangar F, Etemadi A, Keshtkar AA, Hekmatdoost A, et al. Prospective Epidemiological Research Studies in Iran (the PERSIAN Cohort Study): Rationale, Objectives, and Design. American journal of epidemiology 2018;187:647–55.

[11] Eghtesad S, Mohammadi Z, Shayanrad A, Faramarzi E, Joukar F, Hamzeh B, et al. The PERSIAN Cohort: Providing the Evidence Needed for Healthcare Reform. Archives of Iranian medicine 2017;20:691–5.

[12] AT B. Depression: Causes and Treatment. Philadelphia: University of Pennsylvania Press 1972; ISBN 0–8122–1032–8.

[13] Beck AT. Measuring depression: The depression inventory. Recent advances in the psychobiology of the depressive illnesses 1972: 299–302.

[14] Beck AT, Steer RA, Carbin MG. Psychometric properties of the Beck Depression Inventory: Twenty-five years of evaluation. Clinical psychology review 1988;8:77–100.

[15] Gharadingeh K, Manafzade M, Esmkhani R. A SURVEY ON THE AMOUNT OF DEPRESSION IN FEMALES FROM KHOY CITY AND THE FACTORS AFFECTING IT. 2011.

[16] Ghassemzadeh H, Mojtabai R, Karamghadiri N, Ebrahimkhani N. Psychometric properties of a Persian-language version of the Beck Depression Inventory--Second edition: BDI-II-PERSIAN. Depression and anxiety 2005;21:185–92.

[17] Jackson-Koku G. Beck Depression Inventory. Occupational medicine (Oxford, England) 2016;66:174–5.

[18] Lee EH, Lee SJ, Hwang ST, Hong SH, Kim JH. Reliability and Validity of the Beck Depression Inventory-II among Korean Adolescents. Psychiatry investigation 2017;14:30–6.

[19] Yonkers K, Samson JJHoPMW, DC: American Psychiatric Association. MOOD DISORDERS IN. 2000.

[20] Sacco R, Santangelo G, Stamenova S, Bisecco A, Bonavita S, Lavorgna L, et al. Psychometric properties and validity of Beck Depression Inventory II in multiple sclerosis. 2016;23:744–50.

[21] Rossi SR, Greene GW, Rossi JS, Plummer BA, Benisovich SV, Keller S, et al. Validation of decisional balance and situational temptations measures for dietary fat reduction in a large school-based population of adolescents. Eating behaviors 2001;2:1–18.

[22] Lahti J, Ala-Mikkula H, Kajantie E, Haljas K, Eriksson JG, Raikkonen K. Associations Between SelfReported and Objectively Recorded Early Life Stress, FKBP5 Polymorphisms, and Depressive Symptoms in Midlife. Biological psychiatry 2016;80:869–77.

[23] Rohr JK, Xavier Gomez-Olive F, Rosenberg M, Manne-Goehler J, Geldsetzer P, Wagner RG, et al. Performance of self-reported HIV status in determining true HIV status among older adults in rural South Africa: a validation study. J Int AIDS Soc 2017;20:21691.

